# Psychological impact of Covid-19 lockdown in India: Different strokes for different folks

**DOI:** 10.1101/2020.05.25.20111716

**Authors:** Anupam Joya Sharma, Malavika A. Subramanyam

## Abstract

The psychological impact of the lockdown due to the Covid-19 pandemic are widely documented. In India, a family-centric society with a high population density and extreme social stratification, the impact of the lockdown might vary across diverse social groups. However, the patterning in the psychological impact of the lockdown among sexual minorities and persons known to be at higher risk of contracting Covid-19 is not known in the Indian context. We used mixed methods (online survey, n=282 and in-depth interviews, n=14) to investigate whether the psychological impact of the lockdown was different across these groups of Indian adults. We fitted linear and logistic regression models adjusted for sociodemographic covariates. Thematic analysis helped us identify emergent themes in our qualitative narratives. Anxiety was found to be higher among sexual minorities (β=2.44, CI: 0.58, 4.31), high-risk group (β=2.20, CI:0.36, 4.05), and those with history of depression/loneliness (β=3.89, CI:2.34, 5.44). Addiction to pornography was also found to be higher among sexual minorities (β=2.72, CI: 0.09, 5.36). Qualitative findings suggested that sexual minorities likely used pornography and masturbation to cope with the lockdown, given the limited physical access to sexual partners in a society that stigmatizes homosexuality. Moreover, both qualitative and quantitative study findings suggested that greater frequency of calling family members during lockdown could strengthen social relationships and increase social empathy. The study thereby urgently calls for the attention of policymakers to take sensitive and inclusive health decisions for the marginalized and the vulnerable, both during and after the crisis.

## Introduction

The novel coronavirus (Covid-19), taxonomically termed SARS-CoV-2, first emerged in Wuhan, China during late 2019 and was labeled a public health emergency by the World Health Organization (1). The recent and rapid increase in the number of Covid-19 cases, 43,07,287 as on 15^th^ May 2020 globally (2), has increased panic across countries (3). Every country affected by the virus adopted several measures in order to curb its spread. India, home to 1.3 billion people, announced a nationwide “lockdown” on 25^th^ March 2020 (4). The lockdown restricted citizens’ physical mobility, advocated social distancing norms, and limited a majority of public services while allowing the essentials. However, these measures of sheltering-in-place, equivalent to an extended quarantine, likely created a stressful environment for the citizens, given the sudden disruption in their daily routines (5,6). These disruptions could contribute towards adverse psychological outcomes such as post-traumatic stress symptoms (7) and aggressive behaviors (8). For instance, one Indian study by Gautam & Sharma (9), highlighted that lockdown could increase the psychological toll on the Indian academic fraternity because of the disruption in their work, which additionally brings financial instability to the contractual staff. However, the impact of a lockdown might vary across diverse social groups. Individuals who are living alone or away from family (or loved ones), suffering from economic losses, and have a history of negative psychological states, could be at higher risk of depression, loneliness, and anxiety disorders during the lockdown. For instance, queer individuals, known to be already burdened with minority stressors which likely lead to sense of psychological distress (10), could suffer from increased stress during lockdown. Further, restrictions on physical mobility might have not only disrupted their social lives but also paused the sexual lives of many, which could lead to several complications. These disruptions in sexual lives in addition with the on-going stress due to the lockdown could also persuade individuals to (mis)use pornography for coping (11), which could further lead to depressive symptoms (12).

While Covid-19 is non-discriminatory, the psychosocial impact of its spread could also vary across diverse social groups. For instance, the impact on individuals with a known higher risk of contracting Covid-19, such as the elderly or those with co-morbid conditions, might be different from individuals with a lower risk. The extent of daily exposure to the pandemic may also matter: A study from Wuhan, China, found a high prevalence of depressive symptoms among frontline healthcare workers (13). Similarly, another study from Australia showed that people living in high risk infection zones reported higher psychological distress than those living in uninfected areas (14). These findings suggest that a higher perceived risk to Covid-19 could increase anticipatory fear and anxiety. This fear, depression, loneliness, and anxiety during the time of crisis not only could affect mental health but also adversely affect one’s lifestyle and diet, ultimately impacting physical health (15). Previous studies have shown that depression (or anxiety) worsens sleep disorders (16) and eating disorders (17).

Despite these risks, several personal and social resources could be available for individuals to cope with the adverse effects of the crisis. In a family-centric country such as India, family is regarded as a vital social support (18), especially during a crisis. Living with family/relatives (or regular virtual interactions through phone or Internet media) could act as social support which could result in lowering stress during the lockdown. Of note, an opportunity to spend extended time with family members could strengthen family bonds and enhance work-family balance, leading to a better quality of life (19). The lockdown situation has created opportunities for many to spend time with family (and loved ones) and improve the quality of family relationships, if not through physical proximity, virtually. Nevertheless, these opportunities might act as a situational coercion for a few individuals (who have a history of family maladjustment or family conflict) and induce additional stressors, further increasing their vulnerability to adverse psychological outcomes during a lockdown.

In addition to these social resources, several individual-level characteristics such as the nature of employment, access to material resources; and, psychological resources such as resilience-coping and optimism might be beneficial in minimizing the effect of this crisis (20,21). Findings of a recent study from China suggest that positive personal-level characteristics such as emotional-control and optimism could also help minimizing the negative effects of the Covid-19 crisis (22).

Although these concerns warrant attention in the Indian context, we could locate only a few studies reporting the prevalence of depression and anxiety during the Covid-19 crisis in India, including a comparison across age and gender groups (23,24). Moreover, these studies did not examine the prevalence of these outcomes across other social groups, including the vulnerable and the hidden group of sexual minorities. The rapid increase in the number of Covid-19 cases in India and the disruptions due to the lockdown, warrant investigating the processes explaining any social patterning in the psychosocial wellbeing of Indian citizens during this crisis. In response, we conducted a mixed-methods study of Indian adults to unpack how social factors such as sexual orientation, relationship status, and residence in high-infection areas, could be linked with any negative mental health outcomes during the lockdown. We also investigated whether a higher risk of the infection and a history of depression or loneliness worsens the mental health impact of the lockdown. We further explored the complex processes explaining if and how anxiety and depressive symptoms were related to sleeping and eating habits during the lockdown. We also investigated the role individual-level resources played in coping with the effects of the crisis. Because the lockdown likely changed the nature of social interactions, we additionally examined if this brought any change in how individuals viewed the world and increased their social empathy, which could be an important psychological resource for overall wellbeing and quality of life.

### Research questions

The primary research questions explored in this study were:

1. Do levels of adverse psychological outcomes (depressive symptoms, anxiety, addiction to the Internet and pornography, frequency of masturbation, and experiences of hostile attitudes) vary across sexual orientation, relationship status, groups with varying health risk of Covid-19, history of depression/loneliness, and staying in a state with a high number of Covid-19 cases? If yes, what processes explain these differences?
2. Are anxiety and depressive symptoms related to changes in eating and sleeping habits among Indian adults during the lockdown?

In addition to the above primary questions, inspired by the initial two qualitative interviews, we also addressed the following in our study:

3. How are sharing vulnerabilities (stress and depression) with loved ones, and frequency of interaction with family related to strengthening of social bonds and social empathy during the lockdown?

## Methods

### Study design

We followed a convergent mixed methods approach (25,26) in our study. First, two exploratory qualitative in-depth interviews were conducted to refine our research questions. The narratives of the two participants (one male and one female participant) guided us in identifying key factors affecting their mental wellbeing during the time of the lockdown. The participants shared their frustrations related to the lockdown, the disruption in their routine work, the chaos around them regarding the increase in number of cases, their challenges in general, and overall feeling about the entire situation of crisis. Based on the data from these two interviews, we constructed our online quantitative survey. Data for the study were collected through the online survey and qualitative interviews simultaneously. However, whenever the data from the quantitative survey revealed an interesting picture, we dug deeper about it in our qualitative interviews to understand its complexity and context.

## Quantitative survey

We carried out an online survey from 9^th^ May to 15^th^ May 2020. Our survey questionnaire in the form of an anonymous Google form was circulated through several Facebook groups as well as WhatsApp and Instagram contacts. We further used a snowball sampling procedure to increase the number of responses. The authors requested their family members, friends, colleagues, and professional networks to further spread the form among their networks. The introductory passage in the Google form briefed the participants about the broad objective of the study and requested for their voluntary participation. Further, the passage also promised anonymity and confidentiality to the participants. Due to the online nature of the survey, we could not limit its spread to a specific geography. However, we specified our eligibility criteria (Indian citizen, presently residing in India, aged 18 years or above, and who were willing to fill the form in English) in the introductory passage to maximize the chance that we only got responses from India. We received responses from 282 participants.

## Variables and Measures

### Response variables

**Anxiety:** We measured anxiety using the General Anxiety Disorder (GAD-7) scale (27). This widely used scale includes items such as “*Over the past 2 weeks how often have you been bothered by the following problems: Feeling nervous, anxious or on edge?”* The responses were recorded on a 4-point Likert scale ranging from *“not at all* (0)” to *“nearly every day* (3).” We found a good internal consistency of the GAD-7 scale in our sample (Cronbach’s alpha=0.91). The aggregate of the item scores reflected the total anxiety score.

**Depressive symptoms:** We assessed depressive symptoms of our participants using the short version of the CESD-D scale, a 10-item scale (28). The scale includes items such as “In the past week how often have you felt any of these: *I had trouble keeping my mind on what I was doing.”* Two items were reverse scored. The responses to the items varied from *“less than a day”* (0) to *“5-7 days”* (3). We later discovered that responses to one item *(I was bothered by things that usually do not bother me)* did not get recorded possibly due to some technical error in the Google form. However, following Siddiqui (29) and Hawthorne et al. (30), we imputed the person-mean score for the missing item. We found good internal consistency of the scale (including the imputed score) in our sample (Cronbach’s alpha=0.86). The item total was used as the depressive symptom score.

**Symptoms of the Internet addiction:** We used the Internet Addiction Test (IAT) scale (31) to measure symptoms of addiction of the Internet. The scale includes 12 items such as *“Over the past 2 weeks, how often have your felt: find yourself saying “just a few more minutes” when online*?” Responses varied from *rarely* (1) to *always* (5). The Cronbach’s alpha was found to be 0.92. The total score of all 12 items yielded the Internet addiction score.

**Symptoms of pornography addiction:** The Compulsive Pornography Consumption (CPC) scale (32) was used to assess the symptoms of addiction of pornography. The 6-item scale included items such as “*Please indicate how these statements described you during the past 2 weeks: I thought of pornography (porn) when I was trying to focus on other things*.” The responses were recorded on a 7-point Likert scale ranging from *never* (1) to *very frequently* (7). The Cronbach’s alpha was 0.87 in our sample. The sum total of the scores of all items resulted in the pornography addiction score.

**Experiences of hostility:** A single item was used to measure experiences of hostility during the lockdown: *Have you been facing the following problems in the last 2 weeks? You faced a hostile situation (including emotional, physical, and mental violence) from anyone in the place you are currently in*. Responses were recorded as *yes* and *no*.

**Change in food habits (time and consumption):** We assessed any change in the participants’ food habits using a single item: *Have you been facing the following problems in the last 2 weeks? Food patterns (type of foods consumed/timings) have changed*. Responses varied from *not at all* (1) to *always* (5).

**Sleeping problems:** Sleeping problems were measured using a combination of two items, *have you been facing the following problems in the last 2 weeks? Your sleep cycle has changed drastically* and *You have difficulty in falling asleep*. The responses varied from not at all (1) to always (5). The additive score yielded the level of sleeping problem with scores ranging from 2 to 10.

**Frequency of masturbation:** Frequency of masturbation was assessed using a single item, *how often are you engaging yourself in masturbation activities in the last 4 weeks?* The responses varied from *never* (1) to *multiple times a day* (6).

**Social empathy and quality of social relationships:** Social empathy was operationalized based on the participants’ choice of several options offered. The selection of any of the following options: *You have become more socially responsible; you have become more active in neighborhood associations/groups or other social groups near your residence; You have been thinking about the vulnerable in our society and tried to do at least something for them (donating or helping in other ways)* indicated increased social empathy coded as 1 (otherwise 0). Similarly, the quality of social relationships was recorded as 1 (improved), if the participants selected even one of the following options: *You have started liking to spend time with your closed ones more than before; you have strengthened your relationship with your friends; and you have strengthened your relationship with your family/partner*, otherwise as 0.

## Predictors

**Sharing stress and anxiety with loved ones:** If participants selected “yes” to the question *have you shared your stress and vulnerability with loved ones during the lockdown*, it was coded as 1, else as 0.

**Resilience coping:** We assessed resilience coping of the participants using the 4-item Brief Resilience Coping Scale (BRCS) (33). The scale included items such as *I look for creative ways to alter difficult situations*, with responses varying from *does not describe me at all* (1) to *describes me very well* (5). The aggregated score of all items reflected the participants’ resilience. The Cronbach’s alpha was 0.82.

**Optimism:** Optimism was measured using the 10-item Revised Life Orientation Test (LOT-R) scale (34). It included items such as *in uncertain times, I usually expect the best* while responses ranged from, *I disagree a lot* (1) to *I agree a lot* (5). Four items were fillers and were removed from the analysis. The aggregate of all item scores resulted in the optimism score. We found moderate internal consistency of the scale in our sample (Cronbach’s alpha=0.55).

**Change in frequency of calling family members:** We compared the frequency of calling family members during the lockdown with that during October 2019-March 2020. We treated this as an indicator of the change in frequency of calling family members during the lockdown. We coded it as 1 if the frequency increased, otherwise as 0.

**High-risk group:** Individuals who reported having any of the following: chronic respiratory illnesses, diabetes, heart disease, hypertension, or a weakened immune system, were categorized as belonging to the “high-risk group (1)”, else as “low-risk group (0).”

**History of depression/loneliness:** We grouped the participants who reported having a history of depression or loneliness as “group with history of depression/loneliness (1)”, otherwise “group with no history of depression/loneliness (0).”

**Categories of state exposed to Covid-19:** We referred to data from Ministry of Health and Family Welfare, India (35) for categorizing the states as per the counts of Covid-19 cases. We coded Maharashtra (with cases more than 25000 during the data collection) as “highest exposure;” Tamil Nadu, Gujarat and New Delhi (with around 10,000 cases) as “high exposure;” Rajasthan, Madhya Pradesh, and Uttar Pradesh (near to 5000 cases) as “moderate exposure”; and rest of the states as “low exposure.”

**Sociodemographic characteristics:** We also collected information on age (18-29/30-44/45-59/and above 60 years), gender (male/female/others), sexual orientation (straight/queer), relationship status (opposite-sex relationship/same-sex relationship/single/complicated), place of residence (rural/urban), educational qualification (postgraduate/graduate or diploma/12^th^ or lower), and annual income in Indian Rupees (0-3,00,000/3,00,000-10,00,000/10,00,000-20,00,000/above 20,00,000), and the state of residence.

### Qualitative strand

We conducted 14 in-depth interviews from 10^th^ May through 17^th^ May 2020. We circulated an advertisement inviting participants for telephonic interviews through social media (Facebook, Instagram, and Twitter) and personal contacts of the authors. The advertisement included a brief introduction about the study, contact information of the first author (also the interviewer) the nature of the interviews, and about the approximate length of the interview.

The introduction also informed the participants about the sensitive nature of the topic (which included questions on their personal/intimate lives) and asked their preferences of the gender of the interviewer. However, none of the participants shared concerns being interviewed by AJS (a man). Interested participants contacted AJS through email/Facebook/WhatsApp showing their willingness to participate. AJS and the participants mutually agreed on a time for the telephonic interview. Before beginning the interview, AJS once again briefed the participants about the study; and informed them about their anonymity and confidentiality of data, and that the interview would be terminated at any point the participant showed discomfort. In addition to verbal consent, AJS also sought consent to audiotape the interviews. Four participants were reluctant to get the interviews audiotaped. Detailed notes (including several quotes) were taken during these interviews. All interviews began with broader questions like “How do you feel about the entire situation (of Covid-19 and lockdown)?” AJS was cautious while asking personal questions, especially about romantic and sexual lives of the participants. AJS ensured participants’ comfort, not only while asking sensitive questions, but throughout the interview process, by taking a pause and asking, “should we proceed?” However, there were no instances where there arose the need to terminate any interview. All participants shared their emotions, vulnerabilities, moods, challenges, change of lifestyle, and perceived wellbeing during the lockdown (and the Covid-19 crisis). Specific comments regarding the mood and context were noted by AJS to give rigor to the analysis. At the end of the interviews, the participants were requested to share about the study with their peers and network, seeking their participation. The length of the interviews ranged from 28 minutes to 1 hour 40 minutes.

Eight participants were recruited through the advertisement while 2 participants were recruited through snowball sampling. Additionally, we allowed the participants of the online quantitative survey to express interest for a follow-up telephone call. We recruited 4 participants through this method. Different methods of recruitment helped us get a socially diverse sample in a short time.

In addition to the interviews, we collected data through an open-ended question in the online quantitative survey. This allowed the participants to share their concerns related to the pandemic situation (and lockdown). Extracted quotes were used in our qualitative analysis.

## Analyses

### Quantitative analysis

The distribution of all continuous variables was checked for normality (36). Next, we fitted separate multivariable linear regression models to estimate the association of the independent variables (gender, sexual orientation, relationship status, occupation, high-risk group, and living in a states with high number of cases) with psychological outcomes (anxiety, depressive symptoms, Internet and pornography addiction) adjusted for the sociodemographic covariates—age, gender, annual income, educational qualification, place of residence—and for individual personal resources (optimism and resilience). We fitted separate logistic regression models to estimate the associations of sexual orientation and relationship status with the binary variable indicating the experience of hostility, adjusting for all sociodemographic variables.

We also fitted multivariable linear regression models to estimate the association of anxiety and depressive symptoms with changes in sleep and food cycles (separate models), adjusted for the sociodemographic covariates and personal resources.

Additionally, we fitted separate logistic regression models to estimate the association of increased frequency of calling family members with social empathy and the quality of social relationships adjusted for sociodemographic covariates. For all our analyses, alpha was set at 0.05. All statistical models were run in Stata version 12 (37).

We chose a thematic analysis approach (38) to analyze the qualitative data. The analysis began with AJS (who also conducted all the interviews) familiarizing himself with the data by spending prolonged time in re-listening to the audiotaped interviews and reviewing the transcript excerpts. The participants’ narratives about their emotional responses to the situation; their description of how the lockdown affected their routine, relationships, and social responsibilities; their sense of self (including their body); their perspective on life; their coping mechanisms; and, views towards a “new world” guided the coding process. Four themes emerged from these indexed codes (39) and the detailed comments. Additionally, NVT (an external researcher) categorized the themes emerging from the codes. The coding scheme was discussed among the two authors (and NVT), and after critical analysis, the themes were confirmed with a high inter-coder reliability (40). Follow-up interviews with two participants were carried out separately for respondent validation (41). Additionally, several quotes from the open-ended section of the online survey were included in the themes that emerged from the qualitative interviews allowing better representation of all the voices heard.

Both quantitative and qualitative findings carried equal weight in this study. The qualitative themes that emerged gave richer context to the quantitative results during the interpretation phase.

#### Positionality

The study was motivated by previous work of AJS and MAS on the queer community, and their understanding of the community’s unique vulnerabilities. Apart from this, the lockdown has severely restricted the ability of AJS and MAS, not only in terms of physical mobility but also in terms of distance from loved ones and has affected their productivity. Interviews by AJS were conducted with this frame of reference.

### Ethical considerations

The study was approved by the Institutional Ethics Committee, IIT Gandhinagar, India. Utmost precaution was taken by AJS while conducting the telephonic interviews. The participants were informed about the sensitive nature of the questions and were informed that they could skip any question. AJS constantly monitored the mood of the conversation and frequently asked the participants about their willingness to continue. Names of all participants have been changed in this study to protect anonymity.

## Results

### Quantitative results

We analyzed a sample of 282 Indian adults who responded to the online survey. A majority (~75%) of our participants were 30 years or younger. Around 60% identified themselves as male, and about 77% reported to be heterosexuals. Only a small proportion (~12%) of our participants had education less than 12^th^ standard (high school). Greatest proportion (~81%) of the participants resided in Urban areas.

**Table 1:** Sociodemographic characteristics of participants of the quantitative survey (n=282)

|  | Frequency (%) | Anxiety M (SD) | Depressive symptoms M (SD) | Addiction to Internet M (SD) | Addiction to pornography M (SD) |
| --- | --- | --- | --- | --- | --- |
| **Age group (in years)** |  |  |  |  |  |
| 18-29 | 212 (75.71) | 7.07 (5.84) | 12.65 (7.13) | 29.18 (11.53) | 13.43 (8.54) |
| 30-44 | 48 (17.14) | 4.02 (3.68) | 10.21 (7.39) | 22.65 (8.19) | 12.09 (6.58) |
| 45-59 | 15 (5.36) | 6.28 (6.86) | 9.87 (9.05) | 29.55 (16.31) | 11.71 (8.44) |
| 60 and above | 5 (1.79) | 5.8 (6.68) | 11.48 (9.95) | 29 (12.28) | 12.6 (9.04) |
| **Gender** |  |  |  |  |  |
| Male | 175<br>(62.50) | 6.15<br>(5.82) | 11.31<br>(7.36) | 28.43<br>(11.33) | 14.90 (8.37) |
| Female | 102<br>(36.43) | 6.90<br>(5.25) | 13.14<br>(7.06) | 27.43<br>(11.68) | 10.08 (7.08) |
| Other | 3 (1.07) | 12.33<br>(9.07) | 19.62<br>(4.62) | 25.33<br>(13.05) | 11.66 (8.14) |
| **Sexual orientation** |  |  |  |  |  |
| Heterosexual | 218<br>(77.86) | 5.95<br>(5.44) | 11.72<br>(7.39) | 27.26<br>(11.55) | 12.10 (8.02) |
| Sexual minority | 62 (22,14) | 8.23<br>(6.18) | 13.46<br>(6.89) | 30.63<br>(10.75) | 16.44 (8.06) |
| **Educational qualification** |  |  |  |  |  |
| Postgraduate | 160<br>(57.35) | 6.60<br>(5.88) | 12.25<br>(7.52) | 26.99<br>(11.46) | 12.24 (7.53) |
| Graduate/Diploma | 94 (33.69) | 6 (5.44) | 11.78<br>(7.15) | 28.72<br>(10.98) | 14.18 (8.98) |
| 12th or lower | 25 (8.96) | 7.04<br>(5.20) | 12.04<br>(6.72) | 31.14<br>(11.65) | 14.90 (9.55) |
| **Annual income**<br>**(in Indian Rupees)** |  |  |  |  |  |
| 0-3,00,000 | 56 (20.82) | 6.48<br>(5.01) | 13.41<br>(7.41) | 29.22<br>(12.29) | 14.17 (8.13) |
| 3,00,000-10,00,000 | 115<br>(42.75) | 7.27<br>(6.20) | 12.85<br>(6.94) | 29.31<br>(11.55) | 13.65 (8.41) |
| 10,00,000-20,00,000 | 60 (22.30) | 5.93<br>(5.29) | 10.41<br>(6.61) | 26.19<br>(10.14) | 12.45 (8.55) |
| above 20,00,000 | 38 (14.13) | 5.32<br>(5.55) | 10.78<br>(8.12) | 24.19<br>(10.53) | 11.30 (7.39) |
| **Place of residence** |  |  |  |  |  |
| Rural | 53 (18.93) | 6.73<br>(5.52) | 12.48<br>(6.81) | 29.26<br>(11.71) | 12.37 (6.98) |
| Urban | 227<br>(81.07) | 6.42<br>(5.74) | 12.01<br>(7.42) | 27.76<br>(11.39) | 13.24 (8.47) |

### Anxiety and depressive symptoms across social groups

Our fully adjusted models (adjusted for gender, age, educational qualification, income, and place of residence) (see Table 2) found that GAD scores were higher, on average, in sexual minorities (β=2.44, CI: 0.58, 4.31) versus heterosexuals, high-risk group (β=2.20, CI:0.36, 4.05) versus low-risk group, and participants with history of depression/loneliness (β=3.89, CI:2.34, 5.44) versus participants with no history of depression/loneliness. However, GAD scores were lower for single participants (β= -2.35, CI: -4.30, -0.39) than those who were in opposite-sex relationships. Although statistically not significant, our fully adjusted model found lower GAD scores among participants who were in same-sex relationships (β= -0.28, CI: -6.24, 5.68) than opposite-sex relationships. We could not find statistically significant associations of living in a state reporting a high count of Covid-19 cases with anxiety symptoms.

**Table 2:** Associations of sexual orientation, relationship status, risk of Covid-19 infection, history of depression/loneliness, and state’s exposure to Covid-19 (regression coefficients (β), and 95% confidence intervals) with anxiety, depressive symptoms, and addiction to the Internet and pornography (Also sexual orientation and relationship status (adjusted odds rations (AOR, 95% confidence intervals) with experiences of hostility during the lockdown among Indian adults.

|  | Anxiety* | Depressive symptoms* | Addiction to Internet* | Addiction to pornography* | Experiences of hostility (AOR)# |
| --- | --- | --- | --- | --- | --- |
| **Sexual orientation** |  |  |  |  |  |
| Heterosexual (ref) |  |  |  |  |  |
| Sexual minority | 2.440 (0.567, 4.313) | 1.988 (-0.464, 4.441) | 3.638 (-0.075, 7.352) | 2.724 (0.089, 5.358) | 1.632 (0.824, 3.231) |
| **Relationship status** |  |  |  |  |  |
| Opposite-sex relationship (ref) |  |  |  |  |  |
| Same-sex relationship | -0.281 (-6.238, 5.675) | -2.776 (-10.318, 4.766) | -2.701 (-14.234, 8.830) | 9.341 (0.976, 17.705) | 0.529 (0.052, 5.333) |
| Single | -2.347 (-4.306, -0.389) | -2.029 (-4.506, 0.446) | 0.461 (-3.315, 4.238) | 0.541 (-2.168, 3.251) | 0.859 (0.439, 1.680) |
| **Risk to Covid-19** |  |  |  |  |  |
| Low-risk group (ref) |  |  |  |  |  |
| High-risk group | 2.203 (-0.360, 4.046) | 0.832 (-1.565, 3.230) | 0.458 (-3.234, 4.151) | 2.805 (0.223, 5.387) | - |
| **History of depression/loneliness** |  |  |  |  |  |
| No history (ref) |  |  |  |  |  |
| with history | 3.892 (2.343, 5.440) | 4.341 (2.378, 6.304) | 4.547 (1.466, 7.628) | 2.635 (0.406, 4.863) | - |
| **Group of states** |  |  |  |  |  |
| Highest exposure (ref) |  |  |  |  |  |
| High exposure | -1.625 (-4.490, 1.239) | -1.135 (-4.827, 2.557) | -3.488 (-8.992, 2.015) | 0.753 (-3.160, 4.668) | - |
| Moderate exposure | -1.244 (-4.374, 1.885) | -0.086 (-4.072, 3.899) | -2.286 (-8.234, 3.661) | -0.532 (-4.830, 3.766) | - |
| Lowest exposure | -0.972 (-3.885, 1.939) | 0.561 (-3.160, 4.284) | -3.872 (-9.423, 1.677) | 1.491 (-2.485, 5.467) | - |

\* Adjusted for age, gender, income, education, place of residence, optimism, and resilience.

Unsurprisingly, we found a statistically significant association of a history of depression/loneliness with increased depressive symptoms during the lockdown (β=4.34, CI: 2.38, 6.30), independent of other covariates. However, we could not find any evidence linking sexual orientation, relationship status, living in a state reporting a high count of Covid-19 cases, and belonging to a high-risk group with depressive symptoms.

### Addiction to the Internet and pornography, frequency of masturbation across groups

We found that a history of depression/loneliness was statistically significantly associated with higher Internet-addiction symptoms (β=4.55, CI: 1.47, 7.63), independent of all other covariates. However, we could not find evidence that other predictors were associated with Internet addiction.

Moreover, our fully adjusted models showed greater symptoms of pornography addiction, on average, in sexual minorities (β=2.72, CI: 0.09, 5.36) versus heterosexuals, in the high-risk group (β=2.80, CI: 0.22, 5.39) versus low-risk group, in participants in same-sex relationships (β=9.15, CI: 0.93, 17.38) versus opposite-sex relationships, and among those with a history of depression/loneliness (β=2.63, CI: 0.41, 4.86) versus no such history. Additionally, our adjusted models revealed that sexual minorities (β=1.39, CI:0.94, 1.86) and participants in same-sex relationships (β=2.07, CI=0.50, 3.63) reported a higher frequency of masturbation during the lockdown compared to their heterosexual peers.

### Experiences of hostility

We did not find statistical evidence that experiences of hostility differed across sexual orientation and relationship status. Although statistically not significant, sexual minorities (versus heterosexuals) had higher odds of experiencing hostility (AOR=1.63, CI: 0.82, 3.23) during the lockdown independent of the sociodemographic covariates.

### Association of anxiety and depressive symptoms with food and sleep habits

Our fully adjusted models (adjusted for sociodemographic variables and positive resources) (see Table 3) showed that higher depressive symptoms and anxiety symptoms were associated with greater reports of self-reported sleep disorders (β=0.16. CI: 0.11. 0 20 and β=0.19, CI: 0.14, 0.25, respectively) and self-reported changes in food pattern (β=0.05, CI: 411 0.03, 0.08 and β=0.08, CI: 0.05, 0.11).

**Table 3:** Associations of anxiety and depressive symptoms (regression coefficients (β), and 95% confidence intervals) with sleep and food related problems; and of sharing of stress and increased frequency of calling family members (adjusted odds rations (AOR, 95% confidence intervals) with the quality of social relationships and social empathy among Indian adults.

| | Sleep related problems ($\beta$)* | Food related problems ($\beta$)* | Social relationships (quality) (AOR)# | Social empathy (AOR)# |
| --- | --- | --- | --- | --- |
| **Anxiety** | 0.19 (0.14, 0.25) | 0.08 (0.05, 0.11) | - | - |
| **Depressive symptoms** | 0.16 (0.11, 0.20) | 0.05 (0.03, 0.08) | - | - |
| **Sharing of stress with others** | - | - | 2.96 (1.52, 5.74) | 3.99 (1.95, 8.14) |
| **Increased frequency of calling family members** | - | - | 2.56 (1.19, 5.52) | 2.27 (1.06, 4.88) |

\* Adjusted for age, gender, income, education, place of residence, optimism, and resilience.

### Social empathy and quality of relationships

Our fully adjusted logistic regression models (see Table 3) showed that participants who increased the frequency of calling their family members during the lockdown (compared to 6 months earlier) had higher odds of enhancing the quality of their social relationships (AOR=2.56, CI: 1.19, 5.52), and reporting increased social empathy (AOR=2.27, CI: 1.06, 4.88), independent of all sociodemographic covariates. Moreover, our models found that sharing vulnerabilities (stress/depression) with loved ones was associated with higher odds of being socially empathetic (AOR=3.99, CI: 1.95, 8.14), and enhancing social relationships (AOR=2.96, CI: 1.52, 5.74), after accounting for all sociodemographic covariates.

### Qualitative results

Fourteen participants shared with us the slices of their lives during the lockdown. Of the 14 participants, 8 were male, 5 were female, and 1 identified themselves as a non-binary transgender. Six of the 14 identified themselves as sexual minorities. Four were students, 5 worked in private/public sectors (hereafter “service”), and 5 engaged in business/entrepreneurship. The thematic analysis of the narratives of these 14 participants revealed four broad themes. Not all participants’ narratives highlighted all the themes; however, each participant’s narrative was reflected in at least one theme.

#### Theme 1: Emotional responses to *“distance from the real world”*

All 14 participants expressed their unique concerns about the lockdown situation. Words such as “frustrated,” “stressed,” “angry,” and “suffocated,” were frequently used to describe their emotions. Although the intensity of the negative impact of the lockdown varied across participants, most participants (9/10) shared how the lockdown disrupted their lives causing frustration and agitation. For instance, Ashok (name changed) (male, heterosexual, service, in his 20s) shared,

> *Initially it was chill. I thought things would ease out pretty fast. But it now looks like an exceptionally long pause. Each and every day I feel there is an increase of frustration… Hmm… let me put it as… it is like a feeling of anxiety… about the future. We do not know if things would be the same. Especially, am concerned about my work. My work got stuck. Upar se* (on top of that), *you hear about the increasing number of cases in your own neighborhood. All these accumulate and makes you lose your patience. I have not been able to put my focus on work* (since he was working from home)*. And for me, productivity takes very long to revive once it goes down*.

Similar responses of frustration were shared by several students who had mostly enjoyed an outdoorsy life, be it spending time on their college campuses or with friends outside. However, a few shared a different reason for their anxiety: Living in a place with a high number of Covid-19 cases. Rajini’s (female, heterosexual, homemaker, in her 30s) narrative is an example:

> *We are in a containment* (severe movement restriction) *zone, and since the last 12 days none of us have stepped out of the house. And you know what the situation is in Bombay* (Mumbai) *right now! It is pretty stressful […] I have personally kept myself off from news, but there is always social media to give that to you. If you ask me now, I really do not know what the number of cases is, or even what is happening around the neighborhood. I just am desperately waiting for the day when we all will be safe*.

While several respondents shared the fear they felt currently due to a high number of Covid-19 cases in their areas, two respondents, Tulika (female, sexual minority, service, in her 20s) and Salma (transgender (non-binary), sexual minority, service, in their 40s) shared how this “worse” time had forced them to revisit their past trauma. Tulika, who had gone through a break-up one year earlier and was recovering with the help of therapy elaborated,

> *[…] and then sometime in my head it is like a relapse… earlier I would be going out, meeting friends and keep myselfdistracted, but now since you do not have that opportunity, it just keeps coming back […] I have been a workaholic and that’s another thing that in therapy we are trying to work on, but given the ample amount of free time you have; it just keeps coming back to you*.

Another instance of past trauma being triggered was in the case of Salma (they/their/them), who had always managed situations of discrimination (against their transgender identity) calmly but recently lost their temper during such an event.

> *The policewoman stopped us* (them and their partner), *and asked me “what do you think you are,” […] I have always tried to be calm in such situations, but this time, I just lost all my calmness. It was a mix of so many things, my frustration at work during the crisis, my X (a close relative) falling ill just a week before the incident, all these acted together*.

Their stories revealed that the stress and anxiety developed during the lockdown had revived old memories of trauma. Thus, in response, Tulika chose to go through emergency sessions with her therapist, while Salma failed to stay calm and burst out when faced with gender-based discrimination.

While Tulika and Salma revisited trauma, Anurag (male, sexual minority, businessman, in his 30s) who moved to his relatives’ home during the lockdown felt distant from his “real” world. He shared,

> *After I moved out of X* (a city in Uttar Pradesh) *around 17 years ago, this is the longest time I am spending with them* (his close relatives). *They are old and in their 70s, and need me during this time, so I came here to live with them. I came here around the X March, maybe the Y, just before the lockdown. Honestly, I am happy to be here for them, but you know, sometimes it feels like you are so far away from your real world. It took me quite some time to confront my own sexuality And the fact that I stayed away from my X(relatives), who by the way still do not understand the meaning of “gay,” had helped me a lot in coming to terms with myself* (sexuality). […] *I miss meeting people. The longer I am here, the more I feel that I am losing the chance to meet my life partner. I know it may sound silly and selfish to you, but for me at this age, losing* (even) *one single chance of meeting someone is a big thing*.

Anurag’s feeling of distance from his “real” world highlighted how uncomfortable he was at his old home with his relatives. He shared how things around his relatives’ house reminded him of how uneasy he felt while growing up as a gay man. Anurag was conflicted by a dilemma: While the lockdown brought him closer to his relatives at a time of their need, it also placed on him an additional psychological burden.

The suffocation potentially felt by sexual minorities forced due to the lockdown to stay with others to whom they were not out was evident in this quote shared via the online survey:

> *Belonging to sexual or gender minorities in India and spending time with family when you are not open, was already like a cage, with family having lot of expectations and along with societal pressures and humiliations. Closing in* (being confined to) *a non-accepting society and with high-in-expectation family members is destroying my mental health in COVID-19 times. I want to go away from this Society and breathe in fresh air once again where I will not be judged for how I was born*.

While a majority of the participants (9/14) shared mostly about their negative states of mind, a few took a moment to share the positive impact of the lockdown on their lives. For instance, Rumi (female, heterosexual, businessperson, in her 30s) described how she saw the lockdown as an opportunity to introspect about her own life.

> *Not frustrated with it, but I am seeing this as an opportunity that, you know, I have been married for 8 years and there has been so many ups and downs. My life was going without any clarity, but now I am taking this as an opportunity to see what I could do. Life was fast and now it is like I got a break. I do not think of it as a frustration, but it is an opportunity. Since long I was looking for a time where I could sit and do nothing, especially taking a break from my business*.

The narrative of Ashish (male, sexual minority, service, in his 20s. HIV positive) highlighted his equanimity during the crisis. He shared that he did not have any trouble during the lockdown, mostly because he was an introvert who had always loved spending time indoors. However, he mentioned that he and his X (close relative whom he was living with) had been adhering to the usual precautions to avoid the virus. He said that he had always been protective about his health, as was his X (close relative).

#### Theme 2: Impact of the lockdown/Covid-19 on lifestyle

Almost all participants (11/14) described how their daily routines had changed because of the lockdown. While many of them were trying to keep themselves healthy by striving to live a “normal” life, a few mentioned about drastic changes in lifestyle, especially in their eating and sleep patterns. For instance, Tulika shared how she lost motivation to stick to a routine during the lockdown.

> *I usually am a morning person. But ever since this lockdown, my schedule has drastically changed. I have not been waking up early. I am going to sleep around 5 in the morning and continue sleeping till late […] maybe it is because there is no structure anymore. It is more like what do you look forward to. Earlier I used to wake up, looking forward to something. Maybe to go to work and meet people or just to go out. But now I really do not have a point*.

Almost 50% of the narratives suggested such changes in sleep cycles. On the other hand, Subhash (male, sexual minority, service, in his 20s) shared about changes in food habits while also highlighting his mild concerns related to the disruption in his sexual life,

> *I have not been able to keep myself hydrated properly earlier I had a schedule for that too, and now I feel it is really messed*. (He did not have access to a water purifier at home and relied on purchasing mineral water, which was difficult during the lockdown.) *Also, the food, now it is like there is no strict schedule… working out has stopped as well. I guess these are the things that majorly got influenced. Also. sex. Earlier I used to meet people, not very active sexually I would say but 3-4 times a month, but yeah once a while. But now, there is this longer break. […] it just made me hornier that I had been in the normal times. But there is no permanent mental impact honestly. I mean you can watch porn and then masturbate, that is it. But yeah, there is this constant yearning is there in the back of the mind, like, as soon as the lockdown is over, I would start dating again, and so on*.

Rumi however understood this “bad time” as an opportunity to work on herself, a point shared by three other participants. She believed that she could explore a completely different side of hers because of this long break from her busy work. She narrated,

> *I have been experimenting with my life these days. I wake up early and do yoga and then meditation, and then helped my X (a close relative) with some household chores. Currently, I think I feel physically very light, maybe because of exercise. The free time has helped me a lot to explore these, which otherwise was not really possible given the busy life that I had*.

#### Theme 3: Coping with challenges

Each participant shared unique stories of coping with the crisis. While most adapted themselves to the “smaller world,” a few struggled with it and found alternate ways to negotiate their challenges. A few other participants were positive about the crisis, spending time relaxing or pursuing long-held passions. For instance, Rumi described her introspective exploration and enhanced ability to connect with the society through solitude and meditation.

> *[…] then I was doing nothing and looking at trees and people around… and then… cooking has been one thing that I am enjoying these days… I have also been listening to Sadhguru which I have never done before.in fact I have hated all these… but now.maybe I have found him very interesting […] I have also engaged in meditation, and book reading… so I am happy (about this)*.

On the other hand, Tulika kept her mind distracted from the “outside chaos” by immersing herself in social media. For instance, Tulika shared,

> *The number of hours I am awake, I am using social media. Even if I am going to sleep, I will mindlessly keep scrolling until I fall asleep. Because these are the places currently where you see people. Otherwise it is quite just you. I think it is a good place to connect. It keeps me engaged*.

Tulika’s narrative indicates that the online activities have connected her with the outside world, which created an avenue for her to share her vulnerabilities with others through online interactions (messaging and commenting on posts). Notably, more than 50% of the participants reported perceiving the Internet as a way to reduce stress and anxiety during the lockdown. In fact, most of them referred the Internet as the easiest way to keep them distracted from thinking (or overthinking).

A few of the participants who shared their frustration about disruption in their sex lives, reported finding solace in watching pornography and masturbating. Sujoy (male, sexual minority, student, in his 20s) quoted,

> *I think it is the only thing you could right now. I mean I see people online in* Grindr (a dating app for sexual minorities), *and surprisingly people are still looking for sex. And very honestly, I completely understand this desperateness. Most of them, and that includes me, were having our fun days. And all of a sudden this happens. Initially, even I had the thought, ghar ke paas to jake hookup kar sakte hain* (it should be okay to have a hookup close to my place). *But I realized immediately that it is not wise enough to meet people during this time, especially when you know that it* (Covid-19) *could also not show any symptoms[…] What I do right now is watch porn and jerk off*.

Sujoy’s initial inclination to violate the lockdown norms to go out of his home to have sex highlight the repercussions of sudden disruptions in an active sexual life. Similarly, a participant (male, heterosexual, student, 18-25 years old) answering the online quantitative survey shared via the open-ended question:

> *My sexual desires are making me feel more anxious to masturbate, as a single* (man), *very often during this lockdown period*.

#### Theme 4: New perspectives on self, life, and society

Ten of the 14 participants believed that their lives were no longer the same. They believed that they had changed significantly in terms of how they viewed their selves and society in general. For instance, Tulika said,

> *Earlier I used to be worried about very little things, and now it has changed drastically, maybe after spending so much time with myself. I am clearer about my life. I feel I got clarity and I see a huge change in myself. I feel spending time with myself has been the best thing*

Similar observation was made by Rumi, who pointed out that,

> *[…] my mental state is also is very different than before. Earlier I used to get irritated, worried, angry frequently, but now I feel that I am quite easy, surprisingly even with my husband. We used to fight* (giggles) *but now I see that I have been very much calm even with him. It is more because of a lot of things, like about spending time with myself*.

The universal vulnerability, extended time to reflect on their lives, and sharing their vulnerabilities with others had increased the participants’ level of compassion and social empathy, which further strengthened their relationships with their loved ones. This was reflected in what Tulika shared,

> *What I have learnt from this entire situation is that you do not take things for granted anymore, even for like interacting with people. I connected with a lot of friends lately, made a couple of new friends as well, and I feel the conversations are no longer superficial but they seem very real, even though we are not in the same physical space but I feel closer to the people more than when we were closer physically [,..]People have become more vulnerable and have started sharing. Everyone is going through some upheaval right now with this feeling and everyone is trying to connect with others. We are in our mid 20s and everyone is going through this time in pretty much* (a) *similar way, a huge disruption in our lives I would say. So, we become more vulnerable, now I guess these feelings of vulnerability comes out, when you are sharing. And especially when you know that the other person is going through the same as well, and it actually also allows you to connect to the society at large…to a wide range of people*.

Similarly, Rajan (male, heterosexual, student, in his 30s) reflected,

> *Thinking of my parents, who are in their 60s, and knowing about their vulnerability makes me think more and more my own life. Also, you know that you are in a privileged position to understand lockdown and follow self-quarantine measures. But there are people* (who are) *like my parents, in their 60s*, (but unlike his privileged parents) *who cannot afford to sit inside their homes for like 40 days. I have started to think more and more about them, especially knowing that people are dying because of hunger and poverty during this crucial time. While we all are affected by this virus, we all are suffering differently*.

While the narratives of Tulika and Rajan highlighted how their vulnerabilities connected them to the society at large, Rumi shared how the situation (and thus the vulnerability) strengthened her relationship with her husband of eight years. She narrated,

> *I think that I am looking forward to it* (reuniting with her husband who was stuck in a different city due to the lockdown) *and I have a very strong feeling that it would be very different than before, because I am in a completely different situation now. Also, because I was not clear about my life, I would say we did not have a great relationship. I mean not “not great” but we used to fight a lot. But now I feel little concerned and am little worried about him given the whole situation. And then I realized what is important, that the person is important. I also realized where I was wrong and about my frustration. Now I can say I am looking forward to a much better relationship*.

Rumi considered the lockdown situation as an opportunity to reflect upon her own life, tried to connect with people around her, spent longer time in spiritual, motivational, and meditational activities—all of which had helped her find meaning in her life and optimism about her marriage.

## Discussion

Using quantitative data from 282 Indian adults and qualitative narratives of 14 adults, our mixed methods study found that even though the Covid-19 crisis indiscriminately affected everyone, its psychological effects were disproportionate among diverse social groups in India. Our quantitative and qualitative findings both suggest that sexual minority adults, compared to the heterosexuals, are at a higher risk of developing anxiety, depressive symptoms, and addiction to pornography during the lockdown. Moreover, higher levels of anxiety and depressive symptoms were associated with greater disruption in sleep and food cycles. Lastly, our findings unpacked how sharing vulnerability with loved ones, and frequently talking to family members, strengthened social relationships and social empathy among Indian adults during the Covid-19 lockdown.

The higher risk of anxiety in our survey among the sexual minorities than heterosexuals was corroborated by our qualitative findings. Several reasons might explain this. First, previous research suggests that the sexual minority community have a higher prevalence of anxiety and depressive symptoms compared to heterosexuals (42), independent of any crisis. This could be explained using the Meyer’s Minority Stress model (10), which posits that sexual minorities are exposed to unique and additional stressors related to their minority identity, which could combine with other stressors to impact psychological wellbeing. Thus, during the lockdown, their minority stressors (such as sexual orientation-based discrimination and internalized homonegativity) could interact with the lockdown-related stressors, thereby increasing their anxiety much more than that experienced by heterosexuals. Second, the lockdown had likely paused their social as well as sexual lives (which connected them with their own community) which likely restricted their access to a safe space, and limited the social support they received from the community (43). Our qualitative findings corroborate this. Subhash and Sujoy’s narratives showed how an interruption in their active sex life could make the sexual minorities more anxious during the lockdown. This also explains our qualitative finding which suggests a higher likelihood of addiction to pornography and greater frequency of masturbation among sexual minorities (and people in same-sex relationships) during the lockdown compared to heterosexuals (and people in opposite-sex relationships). This disruption in sexual life could explain our quantitative finding which suggested greater anxiety among heterosexuals who were in (opposite-sex) relationships. The lockdown could have resulted in restrictions in physical interactions and romantic dates with their partners, and reduced the social support received, thus increasing their anxiety. Lastly, during the lockdown, it is likely that most adults would move closer to their families for support and to avoid loneliness (44), especially in a family-centric country such as India. However, for many sexual minorities moving in with their parents/relatives, to whom they were not out or who disapproved of their sexuality, could be challenging, increasing their risk of experiencing hostility during the lockdown. Previous studies have shown that parental support and familial environment play crucial roles in self-acceptance among sexual minorities (45,46). The lack of such familial and/or parental support could hinder self-acceptance among sexual minorities, see for instance, Anurag’s narrative of how he could accept his sexuality only after he moved out of his relatives’ home. Moreover, moving away from his “safe space” to a place which brought memories of discomfort likely increased his anxiety. This could be true for several of the sexual minorities who had gone through interpersonal and familial conflict earlier.

Our quantitative findings also found that individuals at greater, versus lower, risk of the effects of Covid-19 showed higher levels of anxiety. A previous study suggested that patients with existing risk factors to Covid-19 such as cardiovascular disease (CVD) were more also more likely have *worse health outcomes* if infected(47). Irrespective of worse health outcomes, belonging to a group with increased risk of contracting Covid-19, which has no known cure and unpredictably causes mortality, could potentially induce additional stress and anxiety. This corroborates findings from previous studies suggesting a higher prevalence of stress among front-line health workers (48), elderly persons (49), and people living with HIV (50,51) during the global public health crisis. However, our qualitative findings are in contrast to this finding. Ashish (who was living with HIV) showed no added concern (or anxiety) due to the lockdown. One explanation for this could be that Ashish was among those who adopt optimism and, in combination with constant precaution, show stronger resilience to adverse situations. Findings from a previous study found that people living with HIV could develop resilience despite their physical and psychological challenges (52). In fact, our study found that optimism and resilience coping were negatively related to anxiety and depressive symptoms. Moreover, Ashish enjoyed spending time indoors, which likely reduced any frustration related to not being able to enjoy regular life, in addition to the lower likelihood of contracting Covid-19.

We did not find quantitative evidence supporting the hypothesis that living in a state with a higher count of Covid-19 cases predicted greater anxiety and depressive in Indian adults. This is in contrast to previous research in Australia which found that respondents living in areas with a high number of influenza cases were at much greater risk of stress than those living in uninfected areas (53). However, our qualitative results supported our hypothesis. The lack of evidence in our quantitative findings could be because of the operationalization of the concept of area. Our study operationalized area-level risk at the state level. It is possible that anxiety was higher among people living in a neighborhood (and not the state) with higher number of Covid-19 cases. Also, in addition to just the count of Covid-19 in the neighborhood (or state) including the infection fatality rate in the operationalization could have given a reliable estimate of the influence of place.

Our quantitative findings suggest that a past history of depression or loneliness could increase anxiety and depressive symptoms during the lockdown. Our qualitative findings corroborate this. Tulika and Salma’s narratives suggest that stress during lockdown could revive past trauma. Previous study findings also support this interpretation (54). Our qualitative and quantitative findings also suggested that increased depressive symptoms in this group could also increase their Internet consumption leading to Internet addiction during the lockdown. Depressed individuals could use the Internet as way to cope with their negative psychological state during the lockdown, which risks addiction during a restrictive state such as a lockdown and could affect their quality of life even after the lockdown.

Anxiety and depressive symptoms during lockdown were found to predict disruptions in sleep and food schedules, corroborating findings from a previous study (55). Qualitative data found that an increase in anxiety and a lack of motivation to lead a routine life increased the risk of an unbalanced sleep cycle, which also impacted food consumption and its timings. Additionally, the inaccessibility to quality food due to the restrictions in physical mobility during the lockdown could affect the balanced diet among Indian adults. Beyond any immediate health effects which could further worsen mental health, such prolonged changes in timing and consumption of food could impact their overall food eating patterns even after the lockdown, resulting in poorer physical and mental health in the longer term as well.

Our quantitative findings suggest that sharing about stress with loved ones and an increase in frequency of interacting with family members likely strengthened social bonds and also increased social empathy among Indian adults. Our qualitative findings elucidate this. Tulika’s narrative highlighted that the universal vulnerability due to the global pandemic and her sharing about it with others in a similar situation improved her connectedness with the people thereby strengthening her social relationships. Similarly, interacting and knowing the vulnerability of Rajan’s parents (more vulnerable) made him more empathetic, and increase his connection with the society at large. This fits with the findings from a previous study that highlighted this argument--sharing and expressing emotions (and vulnerabilities) could make people more empathetic (56). Such increased social empathy could also be a positive response to the pandemic (and lockdown). For instance, a recent study from the West found that higher empathy towards the more vulnerable could induce motivation to maintain and promote social distancing (57).

## Limitations and strengths

There are several limitations of this study that need to be noted while interpreting the results. First, we used psychological scales to measure anxiety, depressive symptoms, Internet addiction, and pornography addiction, instead of clinical interviews which would have yielded medical diagnoses. However, our use of widely cited, reliable, and validated scales are informative, and could indicate symptoms of the psychological outcomes we explore. Second, our recruitment through online media limited our sample to only those who use the Internet, which may have biased the sample towards the higher socioeconomic spectrum. However, the lockdown prevented us from reaching people on the other side of the digital divide. Third, our quantitative study included several shorter and single-item scales (such as the brief resilience coping scale). Longer scales could have yielded robust results. Because our pretest suggested that the length of the questionnaire was perceived as “a lot” we chose to use shorter scales. However, our qualitative findings align with our quantitative findings and unpack the complex processes explaining the psychological outcomes across diverse groups during the lockdown, thereby enhancing our confidence about the conclusions drawn. Lastly, the cross-sectional nature of the study limits our ability to make causal claims. Longitudinal studies with frequent follow-ups during the lockdown could have shed light on causal processes. However, we were grateful to be able to recruit a diverse sample for our quantitative and qualitative strands, which allowed us to explore the differences in the psychological outcomes during the lockdown across different groups in India.

Despite these limitations, our mixed methods findings highlight the additional psychological burden that the lockdown has brought to an invisible group, the sexual minorities. To our knowledge this is the first study to look at the differential psychological impact of the lockdown across different social groups (including sexual orientation) in India. Moreover, our use of qualitative narratives allowed us to understand the processes linking several social factors to the psychological outcomes in a nuanced manner. Our study also highlights a few positive aspects of the lockdown, underscoring the increase in social empathy and strengthened social bonds among Indian adults.

### Implications and conclusions

Our findings echo Balagos’ argument that the marginalization of sexual minorities would be heightened during disasters, because existing inequalities are magnified at such times (58). While the Indian Supreme Court decriminalized homosexual acts in 2018, Indian policies are not yet inclusive of sexual minorities, who remain socially invisible. Our findings call for the attention of counsellors and health professionals in understanding the specific psychological needs of the sexual minorities during such crises and providing services accordingly.

This study highlights the need for regular interaction and emotional support from friends, family, partners, and caregivers of sexual minorities, individuals with a history of depression or loneliness, a higher risk of contracting Covid-19. A recent study (59) highlighted the promise of delivering psychological support through online- and tele-counselling. This study warrants the use of such technologies in an inclusive manner. The study also opens avenues for researchers to further investigate the extent and nature of the psychological impact in such marginalized groups during crises or disasters.

Lastly, our study findings provide evidence for mental health policymakers to begin designing inclusive policies to address the concerns of marginalized groups during and in the aftermath of the Covid-19 global crisis.

All in all, our study highlights the differential psychological effect of the Covid-19 pandemic among sexual minorities, groups with history of depression, and those with high-risk of Covid-19. The study thereby urgently calls for the attention of policymakers to take sensitive and inclusive health decisions for the marginalized and the vulnerable, both during and after the crisis.

## Data Availability

The data cannot be obtained directly because of the sensitive information collected. Please email anupam.sharma@iitgn to obtain the data.

## Acknowledgement

AJS and MAS thank Nilesh Thube (NVT) for his diligent contribution in data management and representation of results. AJS conveys special thanks to Dipankar Dutta, for being extremely patient during the work. AJS and MAS thank Harvansh Dandelia, Rakshit Verma, and several other friends and colleagues who helped in the circulation of the online survey in a short time. AJS thanks his Bula da. AJS thanks all the participants for filling up the form and sharing their emotions during this crisis. AJS was supported by fellowships by Department of Science and Technology, India and IIT Gandhinagar.

